# Safety and immunogenicity of a heterologous booster with an RBD virus-like particle vaccine following two- or three-dose inactivated COVID-19 vaccine

**DOI:** 10.1101/2023.07.17.23292762

**Authors:** Xiaolan Yong, Jun Liu, Ying Zeng, Jing Nie, Xuelian Cui, Tao Wang, Yilin Wang, Yiyong Chen, Wei Kang, Zhonghua Yang, Yan Liu

## Abstract

**Background:** Reduced protection against COVID-19 due to the waning vaccine-induced immunity over time and emergence of immune-evading SARS-CoV-2 variants of concern (VOCs) indicate the need for vaccine boosters. LYB001 is an innovative recombinant SARS-CoV-2 vaccine which displays a repetitive array of the Spike glycoprotein’s receptor binding domain (RBD) on a virus-like particle (VLP) vector to boost the immune system, produced using a Covalink plug-and-display protein binding technology.

**Methods:** The safety and immunogenicity of LYB001 as a heterologous booster at an interval of 6-12 months was assessed in 119 participants receiving a booster with (1) 30μg LYB001 ((I-I-30L) or CoronaVac (I-I-C), (2) escalated dose of 60μg LYB001 (I-I-60L) or CoronaVac in a ratio of 2:1 after two-dose primary series of inactivated COVID-19 vaccine in part 1 of this study, or (3) 30μg LYB001 (I-I-I-30L) after three-dose primary series of inactivated COVID-19 vaccine in part 2 of this study.

**Results:** A well-tolerated reactogenicity profile was observed for LYB001 as a heterologous booster, with adverse reactions predominantly being mild in severity and transient. The peak neutralizing antibody response was observed at 28 days after booster, with GMT (95%CI) against prototype SARS-CoV-2 being 1237.8 (747.2, 2050.6), 554.3 (374.6, 820.2), 181.9 (107.6, 307.6) and 1200.2 (831.5, 1732.3) in the I-I-30L, I-I-60L, I-I-C, and I-I-I-30L groups, respectively. LYB001 also elicited a cross-neutralizing antibody response against the BA.4/5 strain, dominant during the study period, with GMT being 201.1 (102.7, 393.7), 63.0 (35.1, 113.1), 29.2 (16.9, 50.3) and 115.3 (63.9, 208.1) at 28 days after booster in the I-I-30L, I-I-60L, I-I-C, and I-I-I-30L groups, respectively. Additionally, RBD-specific IFN-γ, IL-2, IL-4 secreting T cells, as measured by ELISpot assay, dramatically increased (more than 10 times versus baseline) at 14 days after a single LYB001 booster.

**Conclusions:** Our data confirm the favorable safety and immunogenicity profile of the LYB001 vaccine when used as a heterologous booster, and support the continued clinical development of this promising candidate that utilize VLP platform to provide protection against COVID-19.

**Trial registration:** https://www.clinicaltrials.gov (No. NCT05928455, https://www.clinicaltrials.gov/study/NCT05928455)

## Introduction

SARS-CoV-2 continues to evolve with the emergence of Omicron and its sub-lineages outcompeting other variants of concern (VOCs), resulting in several variant-driven waves of breakthrough infections. Vaccination is the most cost-effective tool to tackle the COVID-19 pandemic. To date, most of the approved vaccines against COVID-19 target the prototype SARS-CoV-2 sequence, including those based on mRNA, adenovirus vector, protein/adjuvant subunit and inactivated virus platforms. These vaccines demonstrated reduced protective effectiveness against COVID-19 over time due to waning immunity and emergence of immune-evading SARS-CoV-2 variants of concern (VOCs). [1–4] In the absence of Omicron-adapted vaccines, optimizing the delivery of first-generation vaccines by using a heterologous booster strategy (mix and match) appeared to induce a better immune response than a homologous booster. [5–10] Two inactivated COVID-19 vaccines (ICVs), developed by Sinovac and Sinopharm from China, accounted for about 45% of global delivered doses in 2021 and notably contributed to worldwide vaccine coverage. [11] These ICVs also proved to be highly effective against severe COVID-19 disease outcomes. [12, 13] However, they exhibited poor or even absent neutralizing antibody (NAb) activity and effectiveness against infection with Omicron sublineages after the two-dose primary series, a primary booster or even a secondary booster. [14–17]

The receptor-binding domain (RBD) on the Spike glycoprotein of SARS-CoV-2 is an immunodominant antigen which contains epitopes for most neutralizing antibodies, [18] and LYB001 is an innovative recombinant vaccine with display of repetitive RBDs on the surface of a virus-like particle (VLP) vector.[19] The array of RBD on the VLP was achieved using a Covalink plug-and-display protein binding technology (isopeptide bond 4T/4C conjunction in Figure 1), similar to platforms described in other research. [20] Because the VLP and RBD can be expressed separately, the modular production of VLP in *Escherichia coli* and RBD in CHO cells is highly scalable. This platform also offers a shortened research and development cycle of a variant-adapted vaccine, if needed, against rapidly evolving pathogens. Vaccine adaptation can thus be easily accomplished, offering a major advantage to tackling major global health challenges in human infectious disease. Additionally, the highly repetitive antigen array (mimicking an actual virus) and relatively large particle size can enhance B cell receptor cross-linking and antigen presenting cell uptake and presentation, leading to strong stimulation of immune cells in the draining lymph nodes and overcoming insufficient immunogenicity that can occur with soluble or monomeric recombinant subunit vaccines. Furthermore, optimal orientation of neutralizing epitope display on the VLP surface can result in higher proportion of neutralizing antibodies. [21] Herein, we therefore present the safety and immunogenicity results of LYB001 used as a booster vaccine at an interval of 6-12 months in two- or three-dose ICV recipients.

**Figure 1:**
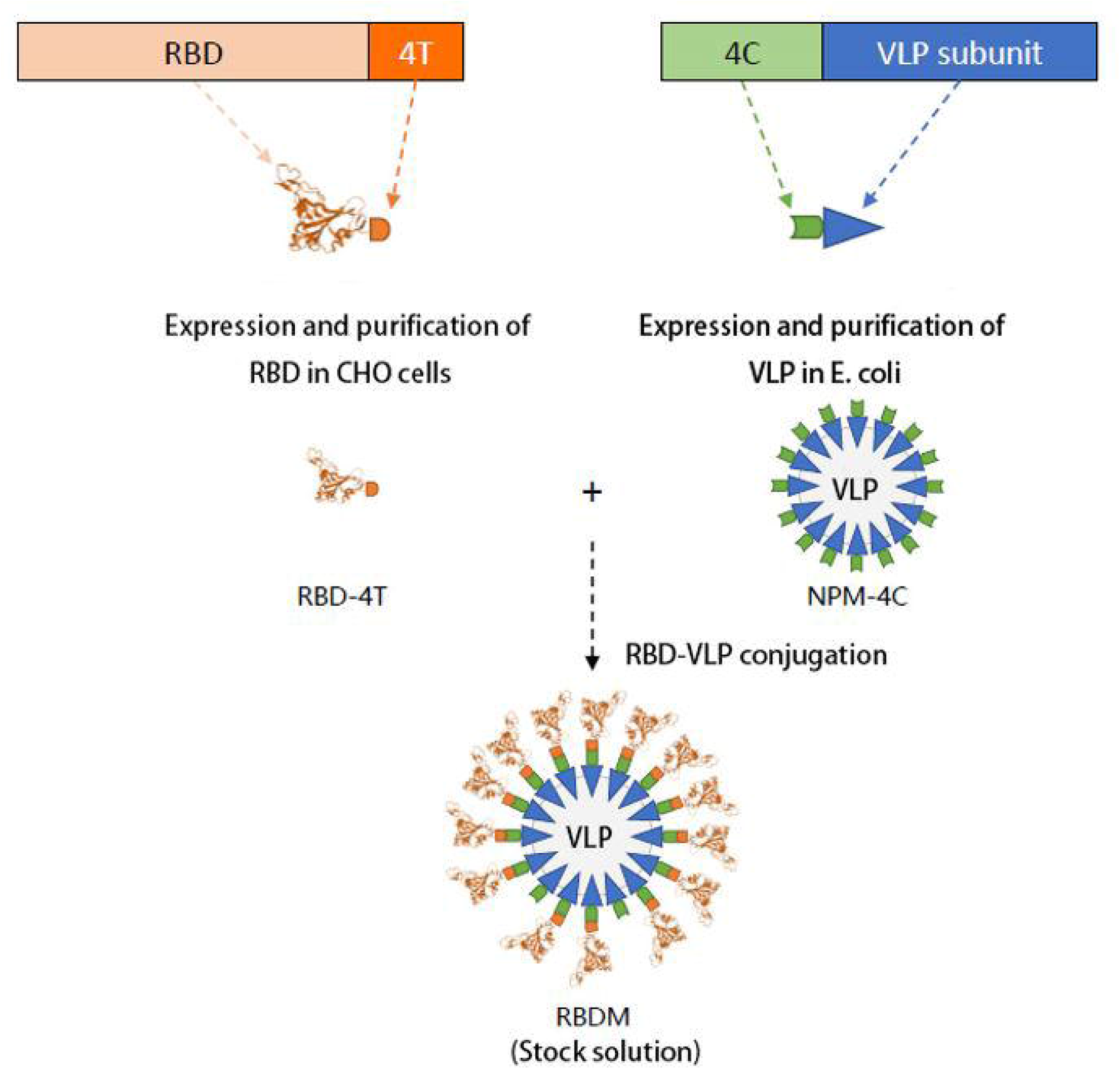
Design principle of LYB001. RBD: receptor binding domain, VLP: virus-like particle

## Methods

### Ethical approval

The study was registered with Clinicaltrilas.gov (NCT05928455), and approved by the Institutional Review Board of Chengdu Xinhua Hospital and Chongqing Red Cross Hospital. The trial was conducted in accordance with the Good Clinical Practice guidelines and Declaration of Helsinki. Written informed consent form from each participant was obtained before any study-related procedures.

### Study design and participants

This study was conducted at Chengdu Xinhua Hospital Affiliated to North Sichuan Medical College and Chongqing Red Cross Hospital (People’s Hospital of Jiangbei District). It was aimed at evaluating the safety and immunogenicity of the heterologous LYB001 booster at an interval of 6-12 months following two or three doses of ICV in healthy participants aged 18-59 years. The study was carried out in two parts: In part 1, a randomized, open-label, positive-controlled design was utilized to evaluate the safety and immunogenicity profile following different heterologous booster doses (30μg and 60μg) of LYB001 using a dose-escalation design. This was compared to a homologous booster dose of CoronaVac in adults 18-59 years of age who had completed a two-dose primary series of ICV 6-12 months earlier. In part 2, a designated dose (30μg) of LYB001 based on the preliminary results from part 1 was used as booster in adults 18-59 years of age who had completed a three-dose primary series of ICV 6-12 months earlier. Participants with a known COVID-19 vaccination history other than ICV, history of SARS-CoV-2 infection, history of severe, uncontrolled chronic disease, or other conditions that, per the judgement of the investigator, might interfere with safety and immunogenicity assessment or pose possible risks to participants, were excluded from the study.

### Randomization and masking

In part 1, the participants were recruited using a dose-escalation study design. Participants who had completed two-dose primary series of ICV were randomly assigned in a ratio of 2:1 either to receive 30μg LYB001 or a CoronaVac booster. After confirmation of an acceptable 7-day safety profile in this cohort, the study was able to proceed to the cohort of two-dose ICV recipients randomly assigned in a ratio of 2:1 either to receive 60μg LYB001 or CoronaVac booster. Randomization of participants and vaccines were performed by an independent statistician using SAS statistical software version 9.4 or higher. Randomization numbers were allocated to eligible participants in the order of enrollment. Participants were randomly allocated to each group in line with the randomization table. In part 2, randomization was not applicable because it was a single-arm study.

Blinding and masking were not applicable for this open-label study as the CoronaVac booster information of each participant had to be mandatorily recorded in the national vaccination system. However, all laboratory staff responsible for immunogenicity assessment and laboratory safety measures were blinded to group allocation.

### Procedures

The design of the investigational vaccine is summarized in Figure 1. Briefly, LYB001 is a recombinant vaccine made by a procedure that expresses VLP vector (NPM-4C) in *Escherichia coli* and RBD (RBD-4T from the Spike glycoprotein of SARS-CoV-2 prototype strain) in CHO cells. Purified stock solutions of RBD-4T and NPM-4C are then mixed to enable conjugation via isopeptide binding to produce the final VLP (RBDM). Following final purification of the VLP, this is finally adsorbed to aluminium hydroxide adjuvant. The LYB001 vaccine was administered through intramuscular injection at doses of 30 or 60μg in a 0.5mL volume. The CoronaVac vaccine was administered through intramuscular injection in a 0.5mL volume.

#### Safety assessments

In this trial, participants were required to stay at the trial site for a 30min safety observation for potential development of immediate adverse events (AEs) after the vaccine booster. During the observation period, participants were instructed to fill out the diary card and given a thermometer and a measurement scale for recording the AEs experienced within 7 days after the booster, including solicited local/systemic and unsolicited AEs. Solicited local AEs included injection-site pain, induration, redness, swelling, rash, and pruritus; solicited systemic AEs included fever, diarrhea, nausea, vomiting, headache, myalgia (non-injection site), cough, fatigue, and acute allergic reaction. On day 8 after booster, participants returned to the trial site for submitting diary cards which were reviewed by the investigator, and contact cards were dispensed to participants for recording unsolicited AEs within 8-28 days after the booster. The intensity of AEs was graded using appropriate guidelines issued by the National Medical Products Administration (NMPA) of China, and the assessment of causality was determined by the investigators.

#### Immunogenicity assessments

Blood samples for humoral immunogenicity assessment were drawn from participants at baseline (day 0 before vaccination), and at days 14, 28 and 90 after booster, and used to determine: (1) Spike glycoprotein binding IgG levels, and (2) NAb titers against prototype SARS-CoV-2 and circulating VOCs. Blood samples for cellular immunity were drawn from the participants at baseline and 14 days after booster. The Spike glycoprotein-binding IgGs were measured using ELISA assays, and values were reported as binding antibody units (BAUs) in accordance with manufacturer’s recommendations and the WHO International Standard and International Reference Panel for anti-SARS-CoV-2 immunoglobulin. The NAb titers against prototype SARS-CoV-2 and dominant VOCs were determined using Vesicular stomatitis virus (VSV)-based pseudovirus neutralizing assays by Chongqing Medleader Bio-Pharm Co., Ltd. Seroconversion was defined as either a four-fold increase in post-boost antibody levels from a seropositive (≥ cutoff value) baseline, or a seropositive conversion from a seronegative (< cutoff value) baseline. The cellular immune response was detected using enzyme-linked immunospot (ELISpot) assay, and was presented as the counts of spot forming cells (SFCs) per 3×10^5^ peripheral blood mononuclear cells (PBMCs) secreting interferon (IFN)-γ, interleukin (IL)-2, IL-4 when stimulated by the RBD peptide pool *ex vivo*. Further details of the methodology used for immunogenicity assays are provided in the *Supplementary Methods*.

### Outcomes

The primary objective of this study was to assess the safety and immunogenicity of LYB001 following a heterologous booster in adults 18-59 years of age who had previously completed a two- or three-dose primary course of ICV vaccination. The primary immunogenicity outcome was the geometric mean titer (GMT), geometric mean fold rise (GMFR) and seroconversion rate (SCR) of spike glycoprotein-binding IgGs, NAb titers against prototype SARS-CoV-2 and circulating VOCs at baseline and 14, 28 days after booster. The primary safety outcome was the immediate AEs within 30 minutes after booster, solicited local/systemic AEs within 7 days and unsolicited AEs within 28 days after booster.

The secondary objective was to assess the immune response durability, which included the GMT and SCR of spike glycoprotein-binding IgGs, NAb titers against prototype SARS-CoV-2 and circulating VOCs measured at 90 days after booster. The secondary safety outcomes were the severe adverse events (SAEs), adverse events of special interest (AESIs) within 90 days after booster, and safety laboratory measures at 3 days after booster. The exploratory outcome was to assess the cellular immune response following a heterologous booster dose of LYB001, and the corresponding exploratory outcome was the RBD-specific IFN-γ, IL-2, IL-4 secreting T cell response as measured by ELISpot assay at baseline and 14 days after booster.

### Statistical analysis

The sample size of this trial was not based on formal statistical hypothesis. Safety analyses were evaluated in the safety set (SS), including all participants who received the booster dose. The immunogenicity analysis was performed in the per protocol set for immunogenicity (I-PPS) following an intention-to-treat principle, including participants who had completed the booster immunization with immunogenicity results at day 0 before vaccination, and at least one available post-boost immunogenicity result with no major protocol deviations. The counts and percentages of participants who experienced AEs were presented in safety analyses, including solicited local/systemic AEs, unsolicited AEs, AEs graded as grade 3 or worse, AEs leading to a participant’s withdrawal, SAEs and AESIs. The NAb GMTs against prototype SARS-CoV-2 and circulating VOCs at different timepoint after booster were calculated with Clopper-Pearson 95% confidence intervals (CIs), and the t-test was used for comparison of log-transformed antibody titers between groups. Additionally, the GMFRs and SCRs at different timepoints after booster, relative to the baseline, were calculated along with their Clopper-Pearson 95% CIs. The cellular immune responses (cytokine secreting T cells by ELISpot assay) and their changes from baseline were statistically analyzed for each group at 14 days after booster, and the differences were statistically tested by Wilcoxon rank-sum test. The χ² test or Fisher’s exact test was used to analyze other categorical data. The statistical analysis was performed using GraphPad Prism 9.0, and *P* < 0.05 was considered statistically significant.

## Results

Between 14 May and 30 June 2022, a total of 215 individuals were screened, and 120 eligible participants aged 18-59 years who competed a two- or three-dose ICV 6-12 months earlier were enrolled in this study (Figure 2). One participant withdrew before booster vaccination due to prior receipt of a two-dose recombinant protein subunit vaccine against COVID-19 (ZF2001) other than ICV. In total, 119 participants aged 18-59 years received the booster vaccination and were included in the analysis set. The mean (SD) participants’ age was 29.4 (8.2), 29.6 (7.8), 30.1 (9.1) and 30.3 (9.4) years in the groups of participants receiving a booster with 30μg LYB001 (I-I-30L), 60μg LYB001 (I-I-60L), or CoronaVac (I-I-C) after two-dose primary series of ICV, or 30μg LYB001 (I-I-I-30L) after three-dose primary series of ICV, respectively. The mean (SD) prime-boost intervals were 290.2 (37.0), 267.8 (31.9), 276.9 (25.0) and 216.4 (37.3) days in the I-I-30L, I-I-60L, I-I-C, and I-I-I-30L groups, respectively. In each group, there were no medication (or vaccination) and medical (or allergic) history which, in the opinion of the investigator, might compromise the participants’ wellbeing, or confound the protocol-specified assessments. The NAb titers against the prototype SARS-CoV-2 and Omicron BA.4/5 variants were low or absent at baseline, and these were comparable across the I-I-30L, I-I-60L, I-I-C groups, and were lower than that of the I-I-I-30L group as anticipated (Table S1).

**Figure 2:**
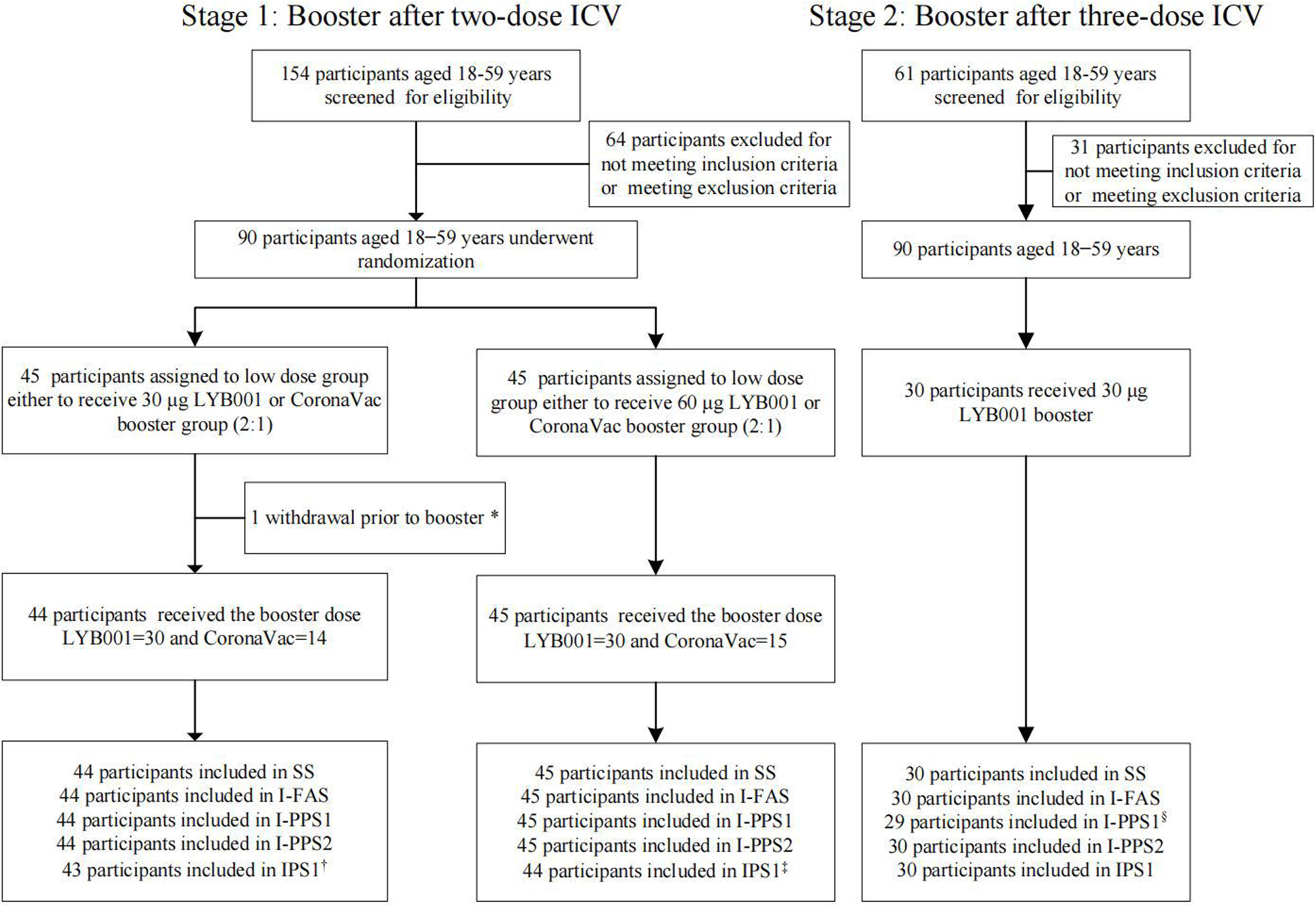
Study profile. *One participant withdrew prior to booster vaccination due to prior receipt of a two-dose recombinant protein subunit vaccine against COVID-19 (ZF2001) other than ICV. ^†^One participant was absent for blood draws at 90 days after CoronaVac booster following two-dose ICV due to quarantine for COVID-19. ^‡^One participant was absent for blood draws at 90 days after CoronaVac booster following two-dose ICV due to quarantine for COVID-19. ^§^One participant was absent for blood draws at 14 days after 30μg LYB001 booster following three-dose ICV due to quarantine for COVID-19. ICV: inactivated COVID-19 vaccine, SS: safety set, I-FAS: full analysis set for immunogenicity, I-PPS: per protocol set for immunogenicity (I-PPS1: I-PPS of 14 days after booster; PPS2: I-PPS of 28 days after booster), IPS: immunogenicity persistence set (IPS1: IPS of 90 days after booster).

The LYB001 as a heterologous booster after a two- or three-dose ICV was safe and well tolerated with adverse reactions (vaccination related AEs) being predominantly mild in severity (only two participants reported grade 2 adverse reactions) (Table 1). The majority of these adverse reactions spontaneously resolved/recovered with a median duration of 2 days after symptom onset and were those commonly anticipated for intramuscularly administered vaccines. The overall incidence rate of adverse reactions was 76.7% (23/30), 66.7% (20/30), 31.0% (9/29) and 63.3% (19/30), which were largely contributed by solicited local adverse reactions, accounting for 70.0% (21/30), 56.7% (17/30), 17.2% (5/29), 60% (18/30) of total participants in the I-I-30L, I-I-60L, I-I-C, and I-I-I-30L groups, respectively. The very common (reported in ≥ 10% of the population in at least one group) solicited local adverse reactions were injection-site pain (predominantly mild in severity), reported in 70.0% (21/30), 56.7% (17/30), 17.2% (5/29), 60% (18/30) participants in the I-I-30L, I-I-60L, I-I-C, and I-I-I-30L groups, respectively. The solicited systemic adverse reactions were reported by 20.0% (5/30), 16.7% (5/30), 3.4% (1/29), 6.7% (2/30) of total participants in the I-I-30L, I-I-60L, I-I-C, and I-I-I-30L groups, respectively. The very common solicited systemic adverse reaction was fatigue, reported in 16.7% (5/30), 10.0% (3/30), 3.4% (1/29), 6.7% (2/30) participants in the I-I-30L, I-I-60L, I-I-C, and I-I-I-30L groups, respectively. The unsolicited adverse reactions within 28 days after booster were reported by 7 (23.3%), 5 (16.7%), 4 (13.8%), and 8 (26.7%), which were comparable across groups. Regarding the safety laboratory measures, the changes at day 3 after booster from baseline did not indicate a particular trend concerning the hematology, blood chemistry, urinalysis, coagulation function parameters, with only a few significantly abnormal safety laboratory parameters being reported (Table 1). All abnormal values spontaneously came back to normal at the subsequent visit without any clinical consequence. There were no SAEs, AESIs, deaths, or AEs that led to withdrawal reported within 90 days after booster. Only one participant in the 60μg LYB001 booster group experienced a grade 3 or worse AE (Preferred terms: Pyrexia) but this was judged as unrelated to the investigational vaccine.

**Table 1.**
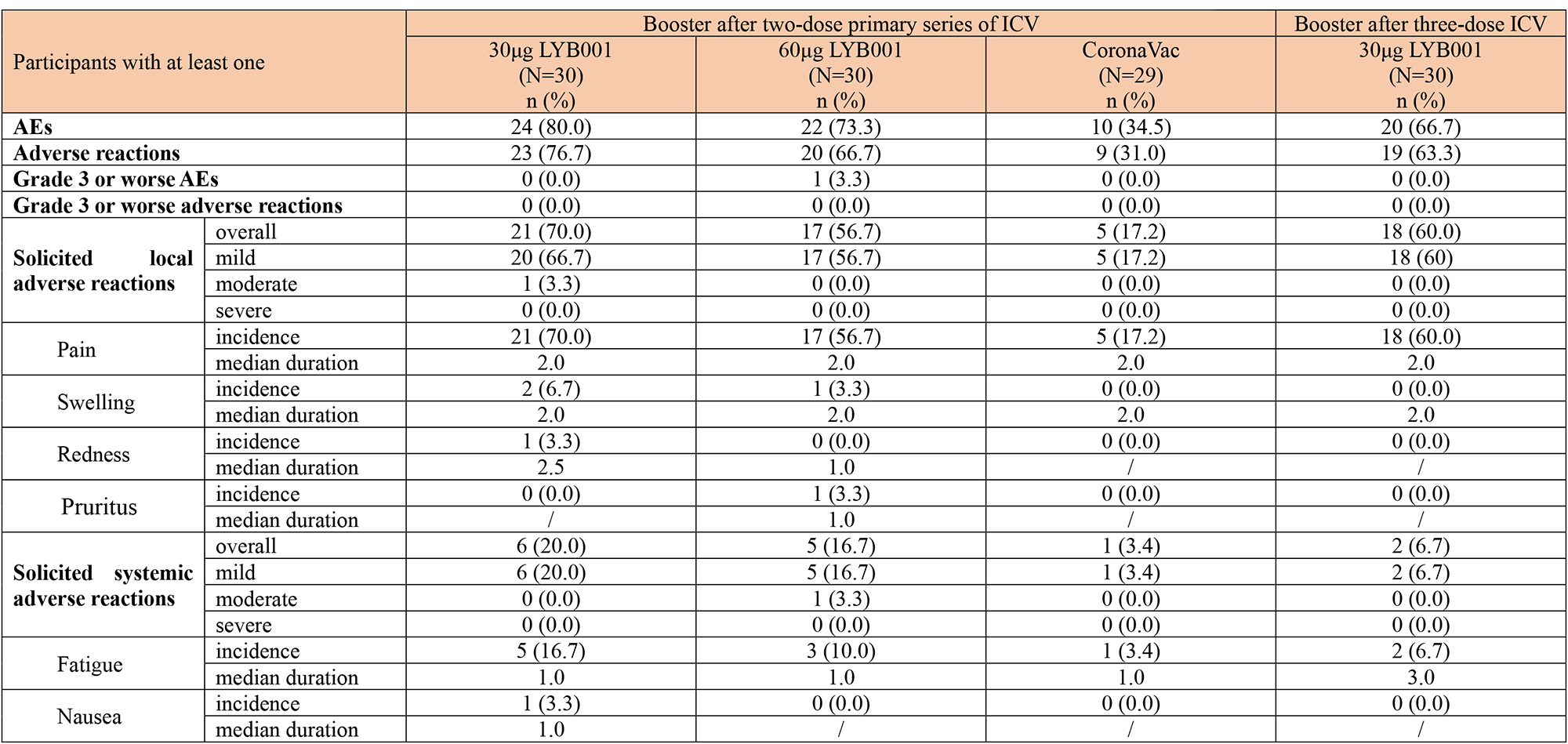
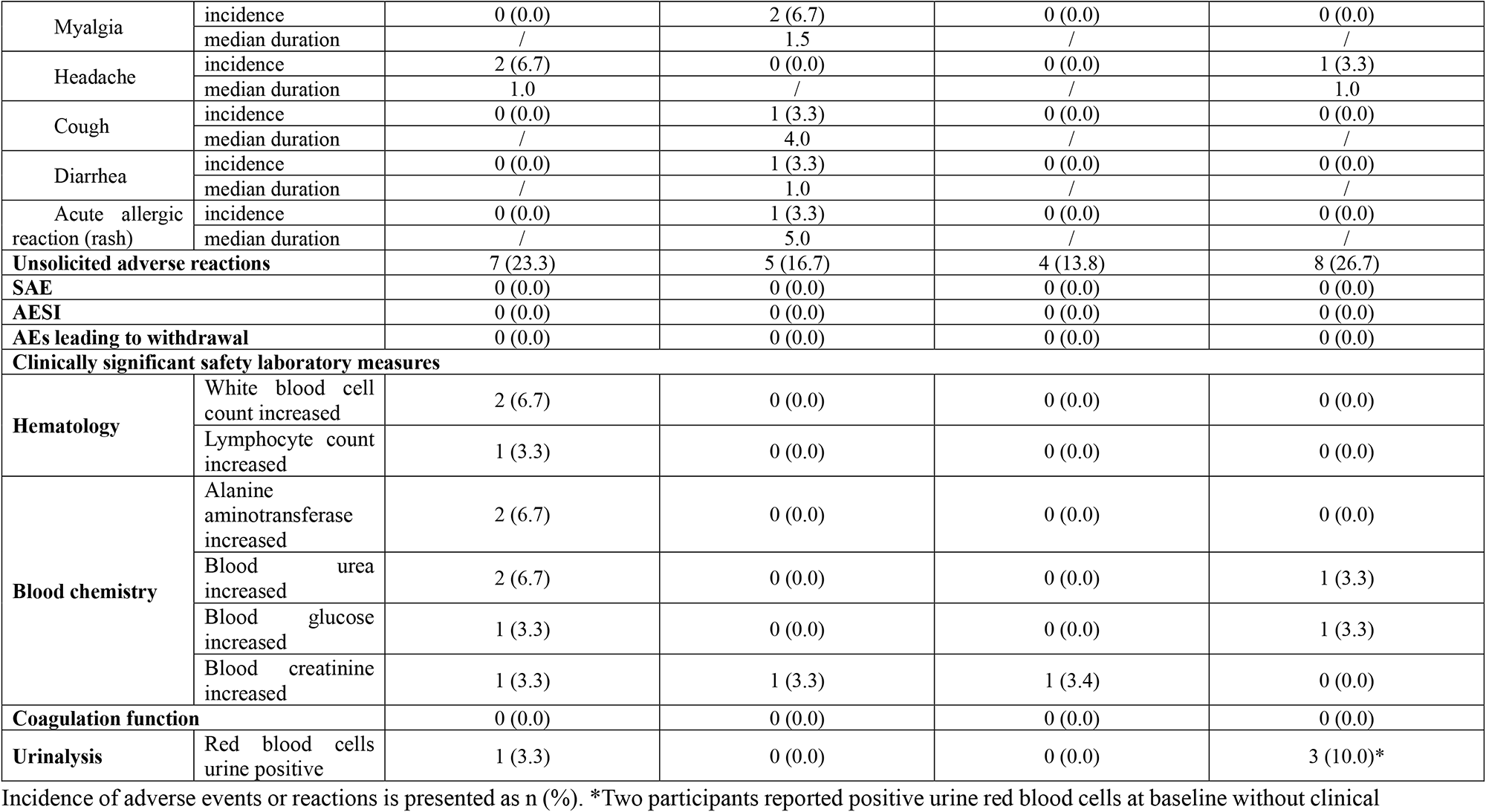
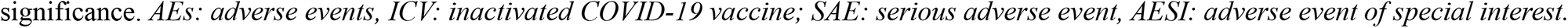
Overall adverse events or reactions after booster.

| Participants with at least one |  | Booster after two-dose primary series of ICV |  |  | Booster after three-dose ICV |
| --- | --- | --- | --- | --- | --- |
|  |  | 30µg LYB001<br>(N=30)<br>n (%) | 60µg LYB001<br>(N=30)<br>n (%) | CoronaVac<br>(N=29)<br>n (%) | 30µg LYB001<br>(N=30)<br>n (%) |
| **AEs** |  | 24 (80.0) | 22 (73.3) | 10 (34.5) | 20 (66.7) |
| **Adverse reactions** |  | 23 (76.7) | 20 (66.7) | 9 (31.0) | 19 (63.3) |
| **Grade 3 or worse AEs** |  | 0 (0.0) | 1 (3.3) | 0 (0.0) | 0 (0.0) |
| **Grade 3 or worse adverse reactions** |  | 0 (0.0) | 0 (0.0) | 0 (0.0) | 0 (0.0) |
| **Solicited local adverse reactions** | overall | 21 (70.0) | 17 (56.7) | 5 (17.2) | 18 (60.0) |
|  | mild | 20 (66.7) | 17 (56.7) | 5 (17.2) | 18 (60) |
|  | moderate | 1 (3.3) | 0 (0.0) | 0 (0.0) | 0 (0.0) |
|  | severe | 0 (0.0) | 0 (0.0) | 0 (0.0) | 0 (0.0) |
| Pain | incidence | 21 (70.0) | 17 (56.7) | 5 (17.2) | 18 (60.0) |
|  | median duration | 2.0 | 2.0 | 2.0 | 2.0 |
| Swelling | incidence | 2 (6.7) | 1 (3.3) | 0 (0.0) | 0 (0.0) |
|  | median duration | 2.0 | 2.0 | 2.0 | 2.0 |
| Redness | incidence | 1 (3.3) | 0 (0.0) | 0 (0.0) | 0 (0.0) |
|  | median duration | 2.5 | 1.0 | / | / |
| Pruritus | incidence | 0 (0.0) | 1 (3.3) | 0 (0.0) | 0 (0.0) |
|  | median duration | / | 1.0 | / | / |
| **Solicited systemic adverse reactions** | overall | 6 (20.0) | 5 (16.7) | 1 (3.4) | 2 (6.7) |
|  | mild | 6 (20.0) | 5 (16.7) | 1 (3.4) | 2 (6.7) |
|  | moderate | 0 (0.0) | 1 (3.3) | 0 (0.0) | 0 (0.0) |
|  | severe | 0 (0.0) | 0 (0.0) | 0 (0.0) | 0 (0.0) |
| Fatigue | incidence | 5 (16.7) | 3 (10.0) | 1 (3.4) | 2 (6.7) |
|  | median duration | 1.0 | 1.0 | 1.0 | 3.0 |
| Nausea | incidence | 1 (3.3) | 0 (0.0) | 0 (0.0) | 0 (0.0) |
|  | median duration | 1.0 | / | / | / |
| Myalgia | incidence | 0 (0.0) | 2 (6.7) | 0 (0.0) | 0 (0.0) |
|  | median duration | / | 1.5 | / | / |
| Headache | incidence | 2 (6.7) | 0 (0.0) | 0 (0.0) | 1 (3.3) |
|  | median duration | 1.0 | / | / | 1.0 |
| Cough | incidence | 0 (0.0) | 1 (3.3) | 0 (0.0) | 0 (0.0) |
|  | median duration | / | 4.0 | / | / |
| Diarrhea | incidence | 0 (0.0) | 1 (3.3) | 0 (0.0) | 0 (0.0) |
|  | median duration | / | 1.0 | / | / |
| Acute allergic reaction (rash) | incidence | 0 (0.0) | 1 (3.3) | 0 (0.0) | 0 (0.0) |
|  | median duration | / | 5.0 | / | / |
| **Unsolicited adverse reactions** |  | 7 (23.3) | 5 (16.7) | 4 (13.8) | 8 (26.7) |
| **SAE** |  | 0 (0.0) | 0 (0.0) | 0 (0.0) | 0 (0.0) |
| **AESI** |  | 0 (0.0) | 0 (0.0) | 0 (0.0) | 0 (0.0) |
| **AEs leading to withdrawal** |  | 0 (0.0) | 0 (0.0) | 0 (0.0) | 0 (0.0) |
| **Clinically significant safety laboratory measures** |  |  |  |  |  |
| **Hematology** | White blood cell count increased | 2 (6.7) | 0 (0.0) | 0 (0.0) | 0 (0.0) |
|  | Lymphocyte count increased | 1 (3.3) | 0 (0.0) | 0 (0.0) | 0 (0.0) |
| **Blood chemistry** | Alanine aminotransferase increased | 2 (6.7) | 0 (0.0) | 0 (0.0) | 0 (0.0) |
|  | Blood urea increased | 2 (6.7) | 0 (0.0) | 0 (0.0) | 1 (3.3) |
|  | Blood glucose increased | 1 (3.3) | 0 (0.0) | 0 (0.0) | 1 (3.3) |
|  | Blood creatinine increased | 1 (3.3) | 1 (3.3) | 1 (3.4) | 0 (0.0) |
| **Coagulation function** |  | 0 (0.0) | 0 (0.0) | 0 (0.0) | 0 (0.0) |
| **Urinalysis** | Red blood cells urine positive | 1 (3.3) | 0 (0.0) | 0 (0.0) | 3 (10.0)* |

Incidence of adverse events or reactions is presented as n (%). \*Two participants reported positive urine red blood cells at baseline without clinical
significance. *AEs*: adverse events, *ICV*: inactivated COVID-19 vaccine; *SAE*: serious adverse event, *AESI*: adverse event of special interest.

As shown in Figure 3 and Table S1, the heterologous LYB001 booster elicited a potent NAb response in participants previously immunized with two- or three-dose ICV, which was low or undetectable at baseline, considerably increased at day 14, peaked at day 28 after booster, and moderately declined by 90 after booster. The VSV-based pseudovirus NAb GMTs (95% CI) against the prototype SARS-COV-2 were 8.5 (6.3, 11.6), 7.0 (5.2, 9.2), 9.0 (6.3, 12.7) and 83.9 (55.5, 126.8) at baseline; 771.6 (452.3, 1316.5), 522.8 (334.5, 817.0), 198.2 (122.5, 320.8) and 1124.4 (727.9, 1737.1) at 14 days after booster; 1237.8 (747.2, 2050.6), 554.3 (374.6, 820.2), 181.9 (107.6, 307.6) and 1200.2 (831.5, 1732.3) at 28 days after booster; 384.3 (232.4, 635.5), 336.8 (215.5, 526.3), 107.7 (62.2, 186.2) and 609.1 (437.3, 848.4) at 90 days after booster, in the I-I-30L, I-I-60L, I-I-C, and I-I-I-30L groups, respectively. The GMFR peaked at 28 after booster, and were 145.0 (85.6, 245.7), 79.8 (53.3, 119.4), 20.3 (11.6, 35.6), and 14.3 (10.2, 20.1) times from baseline in the I-I-30L, I-I-60L, I-I-C, and I-I-I-30L groups, respectively. Despite the decline in NAb response, the SCRs at 90 days after booster remained 100% in the I-I-30L and I-I-60L groups. Versus baseline, the peudovirus NAb titers against prototype at 28 days after booster were 6.8 times higher (P < 0.0001) in the I-I-30L group compared to the I-I-C group. It is also noteworthy that the heterologous LYB001 booster elicited a robust cross-neutralizing antibody response against the Omicron BA.4/5 strain that was dominant during the study period. The pseudovirus NAb GMTs (95% CI) against the Omicron BA.4/5 were 5.0 (5.0, 5.0), 5.0 (5.0, 5.0), 5.0 (5.0, 5.0) and 9.4 (6.2, 14.0) at baseline; 109.6 (52.3, 229.6), 67.3 (36.5, 124.0), 29.9 (17.4, 51.4) and 135.7 (75.6, 243.5) at 14 days after booster; 201.1 (102.7, 393.7), 63.0 (35.1, 113.1), 29.2 (16.9, 50.3) and 115.3 (63.9, 208.1) at 28 days after booster; 44.4 (23.2, 85.2), 32.3 (18.1, 57.7), 14.1 (8.8, 22.5) and 42.0 (24.7, 71.4) at 90 days after booster, in the I-I-30L, I-I-60L, I-I-C, and I-I-I-30L groups, respectively. The NAb against BA.4/5 at 28 days after booster was 6.9 times higher (P < 0.0001) in the I-I-30L group compared to the I-I-C group from an equivalent baseline. The spike glycoprotein binding IgGs exhibited a similar trend to those seen for NAb responses, but with a slower waning at 90 days after booster (Figure S1).

**Figure 3.**
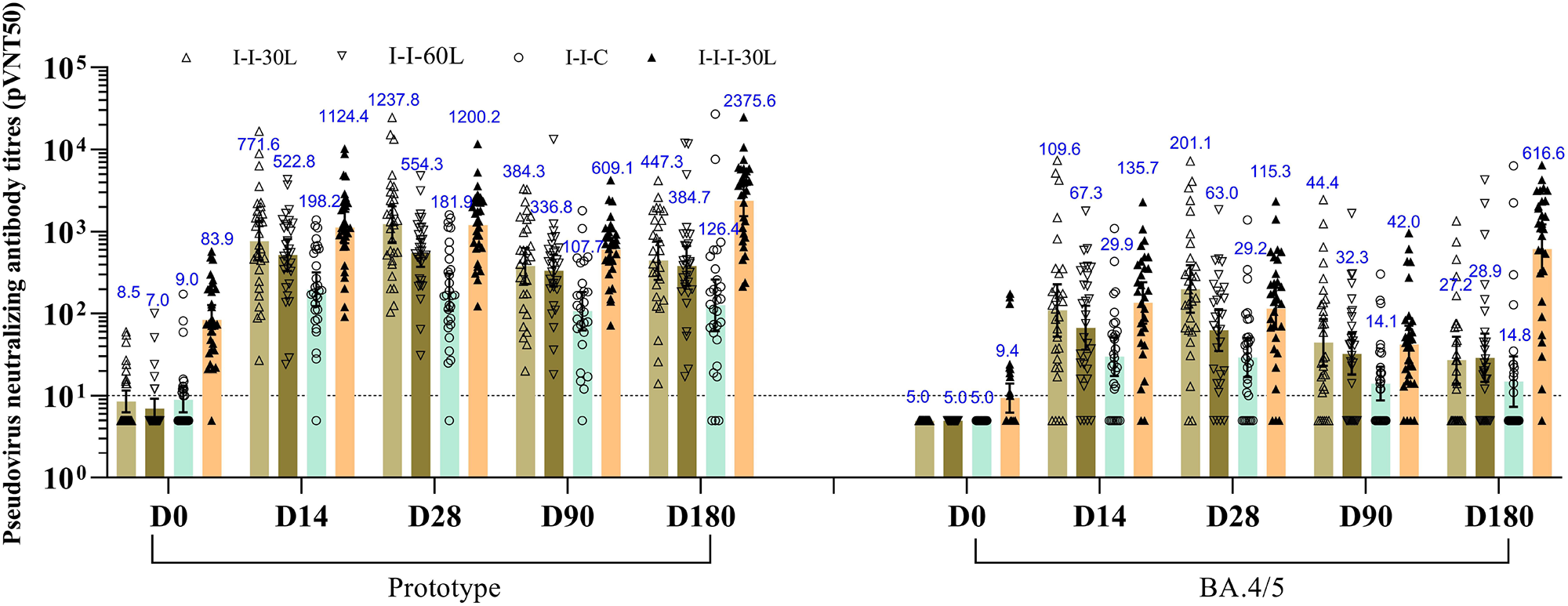
The VSV-based neutralizing antibody titers against prototype SARS-CoV-2, Omicron BA.4/5 strain. Antibody values reported as below the lower limit of detection (LOD=10) were replaced by 0.5*LOD. The individual data in the I-I-30L, I-I-60L, I-I-C, and I-I-I-30L groups are indicated by △, ▽, ○, ▴, respectively. VSV: Vesicular stomatitis virus, I-I-30L: 30μg LYB001 booster after two-dose inactivated COVID-19 vaccine, I-I-60L: 60μg LYB001 booster after two-dose inactivated COVID-19 vaccine, I-I-C: CoronaVac booster after two-dose inactivated COVID-19 vaccine, I-I-I-30L: 30μg LYB001 booster after three-dose inactivated COVID-19 vaccine.

As shown in Table S2 and Figure 4, the LYB001 booster induced significantly higher cytokine responses to the SARS-CoV-2 RBD peptide pool both in the 30μg LYB001 and 60μg LYB001 groups as compared to the CoronaVac group (*P* < 0.001). In most treatment groups, a minority of participants in each group had relatively low pre-existing IFN-γ, IL-2, IL-4 responses to the RBD peptide pool, while the majority of participants exhibited pre-existing responses close to or below the LOD. For IFN-γ, SFCs per 3×10^5^ PBMCs as indicated by median (Q1, Q3) were 1.0 (0.0, 2.0), 0.0 (0.0, 1.0), 0.0 (0.0, 2.0) and 0.0 (0.0, 1.0) at baseline, and 23.0 (8.0, 68.0), 23.0 (10.0, 42.0), 2.0 (0.0, 4.0) and 18.0 (5.0, 46.0) at 14 days after booster in the I-I-30L, I-I-60L, I-I-C, and I-I-I-30L groups, respectively. For IL-2, the SFCs per 3×10^5^ PBMCs as indicated by median (Q1, Q3) were 4.0 (1.0, 10.0), 2.5 (0.0, 5.0), 5.0 (2.0, 8.0) and 0.0 (0.0, 3.0) at baseline, and 48.0 (26.0, 145.0), 39.0 (21.0, 70.0), 5.0 (3.0, 8.0) and 54.0 (30.0, 99.0) at 14 days after booster in the I-I-30L, I-I-60L, I-I-C, and I-I-I-30L groups, respectively. For IL-4, the SFCs per 3×10^5^ PBMCs as indicated by median (Q1, Q3) were 1.0 (0.0, 3.0), 0.0 (0.0, 1.0), 1.0 (0.0, 1.0) and 0.0 (0.0, 2.0) at baseline, and 12.0 (4.0, 36.0), 8.0 (4.0, 33.0), 1.0 (0.0, 1.0) and 18.0 (7.0, 43.0) at 14 days after booster in the I-I-30L, I-I-60L, I-I-C, and I-I-I-30L groups, respectively. While the cellular response measured in I-I-C group was almost absent, both the 30μg and 60μg LYB001 booster induced a balanced Th1/Th2 type cellular response, exhibiting over ten-fold increases versus the respective median SFCs at baseline.

**Figure 4.**
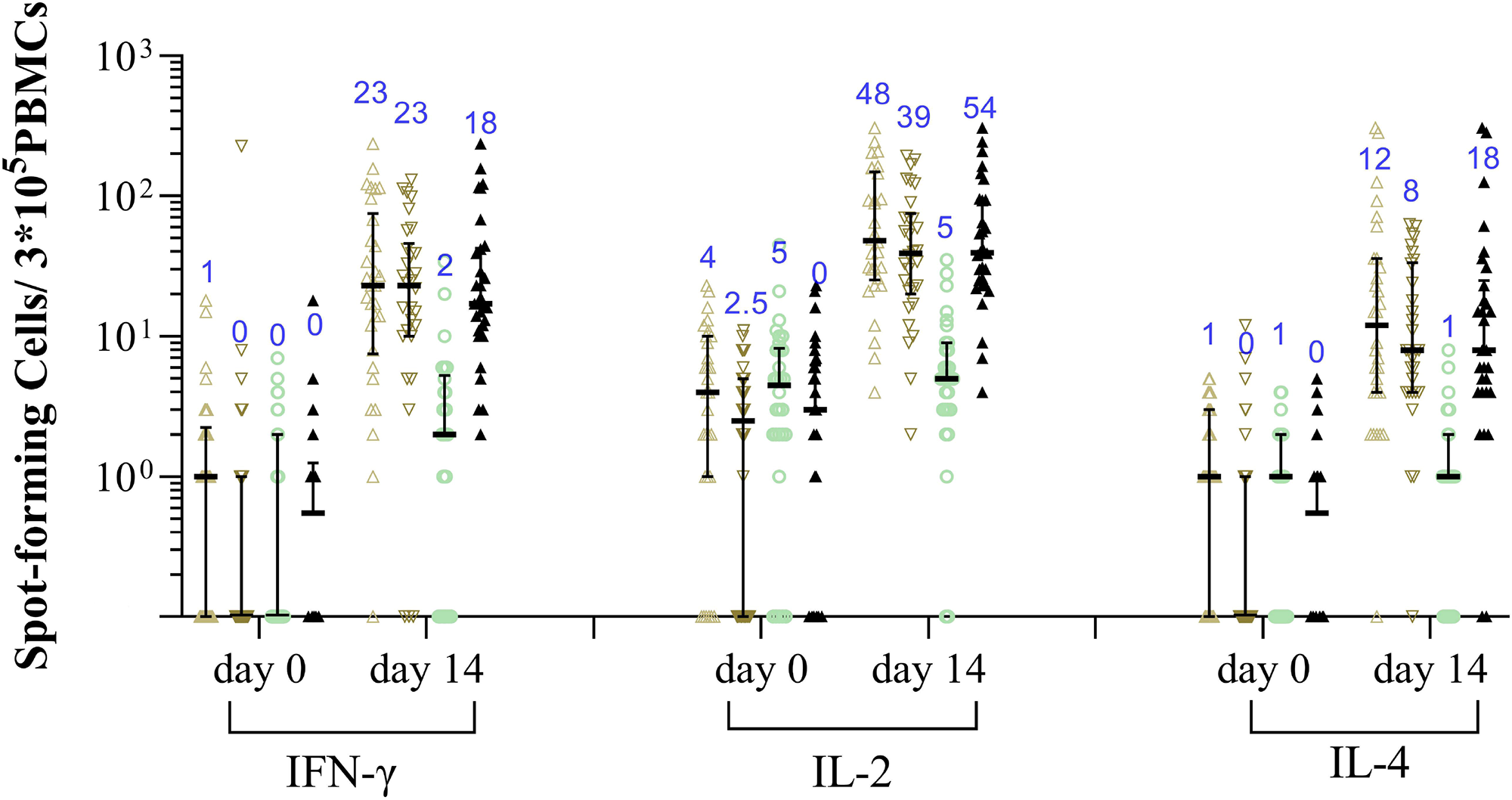
RBD-specific IFN-γ, IL-2 or IL-4 secreting T cells measured by ELISpot assay. Bars and numbers in the figure indicate group medians, and error bars indicate interquartile range. The individual data in the I-I-30L, I-I-60L, I-I-C, and I-I-I-30L groups are indicated by △, ▽, ○,, ▴, respectively. RBD: receptor binding domain, I-I-30L: 30μg LYB001 booster after two-dose inactivated COVID-19 vaccine, I-I-60L: 60μg LYB001 booster after two-dose inactivated COVID-19 vaccine, I-I-C: CoronaVac booster after two-dose inactivated COVID-19 vaccine, I-I-I-30L: 30μg LYB001 booster after three-dose inactivated COVID-19 vaccine.

## Discussion

In this study, we investigated the safety and immunogenicity of LYB001 as a heterologous booster following two or three doses of ICV in participants aged 18-59 years at a prime-boost interval of about 6-12 months. To the best of our knowledge, this is the first clinical trial in China in which the preliminary safety, reactogenicity and immunogenicity results of a VLP-based vaccine against COVID-19 are reported. Overall, despite higher incidence of reactogenicity events compared to the I-I-C group, AEs were mostly mild in severity and transient after a heterologous booster with LYB001 across the I-I-30L, I-I-60L, and I-I-I-30L groups. The reported solicited adverse reactions appear to represent reactogenicity events anticipated for intramuscularly administered vaccines, with local AEs like injection-site pain, swelling, redness, pruritus, and systemic AEs like fatigue. In addition to the mild reactogenicity, the majority of AE symptoms resolved spontaneously, mostly within 48h after onset, and those requiring treatment were managed with simple measures and widely available medications. No vaccination-related SAEs, AESIs, or AEs leading to the participant’s withdrawal were reported within 90 days after booster. Although some participants reported abnormal safety laboratory measures with clinical significance at 3 days after booster, abnormal values spontaneously came back to normal at the subsequent visit without any clinical consequence, and the differences from baseline (calculated by values at day 3 after booster minus baseline values) for each participant in the I-I-30L, I-I-60L, I-I-C, and I-I-I-30L groups did not indicate a particular trend. After further analysis, we found that the higher incidence rate of solicited local/systemic for heterologous LYB001 booster (in I-I-30L, I-I-60L, and I-I-I-30L groups) compared to CoronaVac booster (in I-I-C group) was predominantly contributed by injection-site pain, accounting for 70.0% (n=21), 56.7% (n=17), 60.0 (n=18), versus 17.2% (n=5) of the participants in the I-I-30L, I-I-60L, I-I-I-30L groups, versus the I-I-C group, respectively. The possible explanations for the increased incidence of injection-site pain are: (1) Aluminum adjuvant content in LYB001 was higher than that of the inactivated vaccine (higher aluminum adjuvant was previously reported to correlate with higher risk of pain), [22] or (2) LYB001, with display of repetitive RBD on VLP vector, has larger particle size, which may result in longer local recruitment time of relevant molecules of the innate immune system and activation of antigen-presenting cells. [21] Similarly, mRNA vaccines elicit transient increases in C-reactive protein (CRP), which is an indicator of vaccine adjuvant activity. [23] Our results were also consistent with a self-assembling, two-component nanoparticle vaccine (approved in South Korea) displaying the RBD of the SARS-CoV-2 Spike glycoprotein in a highly immunogenic array. This vaccine demonstrated injection-site pain in 88.1% of participants receiving 10μg GBP510 and 92.3% of participants receiving 25μg GBP510 in AS03 adjuvant. [24] In a phase Ⅲ trial of another coronavirus-like particle (CoVLP) vaccine, injection site pain was reported in 85.0% of participants after the first dose of CoVLP with AS01 adjuvant versus 29.4% of participants in the placebo group.[25]

The results from our study indicate that one heterologous booster dose of LYB001 can profoundly restore the NAb response irrespective of the baseline antibody levels. The LYB001 elicited a comparable NAb response (1237.8 vs 1200.2) against the prototype at 28 days after booster in I-I-30L group versus I-I-I-30L group, despite a significantly different baseline (8.5 vs 83.9, *P*<0.05). The heterologous LYB001 booster also induced superior humoral immune responses in comparison to the homologous booster with CoronaVac. When compared with a recombinant fusion protein vaccine V-01 (IFN-PADRE-RBD-Fc dimer, already approved in China), the peak geometric mean ratio (GMR, heterologous investigational vaccine booster versus homologous ICV booster) regarding VSV-based pseudovirus NAb GMT against prototype in the 30μg LYB001 group was 6.8 (1237.8 vs 181.9), higher than the GMR of 3.3 (893 vs 268) after 10μg V-01 booster. The peak GMR (heterologous booster versus homologous ICV booster) was 6.9 (201.1 vs 29.2) regarding the VSV-based pseudovirus NAb GMT against Omicron BA.4/5, higher than the GMR of 3.8 (211 vs 56) against BA.1 after V-01 booster. [26] Additionally, for a recombinant protein vaccine ZF2001 (already approved in China, Colombia, Indonesia, Uzbekistan), the GMR (25μg ZF2001 booster versus homologous ICV booster) was 2.4 (537 vs 225) regarding the VSV-based pseudovirus NAb GMTs against prototype SARS-CoV-2, and 1.7 (108 vs 63) times regarding the pseudovirus NAb GMTs against Omicron BA.1 of that after ICV booster. [27] Taking into consideration a similar NAb detecting technique (VSV-based pseudovirus NAb assay), trial population (participants aged 18-59 years who completed a two-dose primary series of ICV) and comparison based on the active comparator of ICV, the 30μg LYB001 booster appeared to elicit a more favourable neutralizing antibody response against the circulating variant during the study period as compared to the two approved recombinant protein subunit vaccines (V-01 and ZF2001).

Albeit the LYB001 was designed using the RBD from the prototype SARS-CoV-2, it demonstrates satisfactory immunogenicity against the prototype SARS-CoV-2 and robust cross-neutralizing activity against Omicron BA.4/5-SARS-CoV-2 variants that showed extensive immune escape. The conserved neutralizing epitopes between Omicron BA.4/5 and prototype SARS-CoV-2 might contribute to the cross neutralization. The immunogenicity could be also augmented by the innovative platform using the RBD-VLP protein binding technology, which enhance B cell activation, APC uptake and presentation, and efficient draining to lymph nodes. The optimal orientation for maximizing neutralizing epitope display, leading to a higher proportion of functional antibody, is also reflected by a higher fold rise with regard to the ratio of NAb titer versus Spike glycoprotein binding antibody concentration when comparing LYB001 booster with CoronaVac booster. Thus, the innovative design enables LYB001 to elicit robust neutralizing antibody responses against prototype SARS-CoV-2 as well as cross-neutralizing activity against circulating Omicron BA.4/5. Although a correlate of protection has not been established for predicting individual-level risk of SARS-CoV-2 infection, a spike glycoprotein binding IgG concentration of 1148 BAU/mL (also reported as BAUs in accordance with the WHO Standard) may provide 75% protection against symptomatic infection with BA.5, [28] indicative of a promising efficacy against BA.5 after LYB001 booster.

T cell responses were also important in controlling disease development in patients with COVID-19. Targeted T cell epitopes were broadly conserved between prototype SARS-CoV-2 variant and Omicron. [29, 30] Generally, the cellular immune response is absent or at least weak after booster in healthy adults receiving two-dose ICV, in consistent with previous findings. [31, 32] The results from this trial indicated that the LYB001 booster induced robust cellular responses to the SARS-CoV-2 RBD-specific peptide pool in the I-I-30L, I-I-60L, and I-I-I-30L groups, as compared to the absent cellular responses in I-I-C group. The RBD-specific IFN-γ secreting T cells measured by ELISpot assay were dramatically increased (more than 10 times versus baseline) after a single LYB001 booster, comparable to one shot of adenovirus type-5-vectored COVID-19 vaccine (a median of about 10 IFN-γ secreting SFCs per 1×10^5^ PBMCs) which generally elicited robust cellular immune response. [33] A proportion of the participants in this study appeared to have pre-existing cellular responses to the RBD-specific peptide pool used for PBMC re-stimulation. Such cross-reactive T cell memory was possibly due to previous exposure to common human coronaviruses. [34, 35]

This study also has limitations. First, the safety and immunogenicity findings were concluded based on a small sample size (about 30 in each group), so the results should be interpreted with caution. Besides, our study is an open label study (the participants receiving the CoronaVac booster must be registered with the vaccination system of China, and the fourth dose of CoronaVac booster, which was used for active comparator in part 1, was not approved when the study began), so the evidence provided in this trial is weaker than in a blinded randomized control trial (RCT). The blinded RCTs of LYB001 booster with larger sample size and with age stratification (including older participants aged ≥ 60 years) have now been conducted. Additionally, we only detected the NAb titers of emerging Omicron sublineages during our study period in China, e.g., BA.5. Other emerging VOCs, such as XBB and its sublineages, will be included in our further studies.

## Conclusions

Taken together, the LYB001 as a heterologous booster, after inactivated COVID-19 vaccine, elicited a strong humoral immune response, demonstrated cross-neutralizing activity against dominantly circulating BA.4/5 variants and induced robust RBD-specific cytokine secreting T cell responses, without compromising vaccine’s safety. The reactogenicity events were predominantly mild (grade 1) in severity and transient, and were anticipated for intramuscularly administered vaccines. Thus, our study provides valuable evidence for the LYB001 vaccine, developed using the RBD-VLP protein binding technology, as a promising candidate for preventing COVID-19.

## Funding

This work was partially supported by a grant from the Key Program of Yantai Science & Technology Bureau (No. 2022CXG010508). The study was funded by Patronus Biotech Co., Ltd.

## Data Availability

The data used in the current study are available from the corresponding authors upon reasonable request.

## Conflict of Interests

Ying Zeng, Wei Kang, and Zhonghua Yang are employees of Yantai Patronus Biotech Co., Ltd. The other authors declare no competing interest.

## Author Contributions

All authors have read and approved the article. Yan Liu, Xiaolan Yong, Jun Liu, Ying Zeng, Xuelian Cui and Yilin Wang conceived and designed the clinical study. Xiaolan Yong, Jun Liu and Jing Nie performed the clinical trial. Tao Wang performed the immunological (humoral and cellular) assays. Yiyong Chen, Wei Kang and Yan Liu were engaged in manuscript preparation, manuscript editing. Zhonghua Yang reviewed data analysis. All authors reviewed the manuscript.

## Notes

### Clinical Trial

NCT05928455

### Author Declarations

The study was approved by the Institutional Review Board of Chengdu Xinhua Hospital and Chongqing Red Cross Hospital.

## References

1. Goldberg Y, Mandel M, Bar-On YM, Bodenheimer O, Freedman L, Haas EJ, et al. Waning Immunity after the BNT162b2 Vaccine in Israel. N Engl J Med 2021;385:e85. doi: 10.1056/NEJMoa2114228.

2. Andrews N, Stowe J, Kirsebom F, Toffa S, Rickeard T, Gallagher E, et al. Covid-19 Vaccine Effectiveness against the Omicron (B.1.1.529) Variant. N Engl J Med 2022;386:1532–1546. doi: 10.1056/NEJMoa2119451.

3. Abu-Raddad LJ, Chemaitelly H, Bertollini R, National Study Group for C-V. Waning mRNA-1273 Vaccine Effectiveness against SARS-CoV-2 Infection in Qatar. N Engl J Med 2022;386:1091–1093. doi: 10.1056/NEJMc2119432.

4. Zeng G, Wu Q, Pan H, Li M, Yang J, Wang L, et al. Immunogenicity and safety of a third dose of CoronaVac, and immune persistence of a two-dose schedule, in healthy adults: interim results from two single-centre, double-blind, randomised, placebo-controlled phase 2 clinical trials. Lancet Infect Dis 2022;22:483–495. doi: 10.1016/S1473-3099(21)00681-2.

5. Li J, Hou L, Guo X, Jin P, Wu S, Zhu J, et al. Heterologous AD5-nCOV plus CoronaVac versus homologous CoronaVac vaccination: a randomized phase 4 trial. Nat Med 2022;28:401–409. doi: 10.1038/s41591-021-01677-z.

6. Munro APS, Janani L, Cornelius V, Aley PK, Babbage G, Baxter D, et al. Safety and immunogenicity of seven COVID-19 vaccines as a third dose (booster) following two doses of ChAdOx1 nCov-19 or BNT162b2 in the UK (COV-BOOST): a blinded, multicentre, randomised, controlled, phase 2 trial. Lancet 2021;398:2258–2276. doi: 10.1016/S0140-6736(21)02717-3.

7. Callaway E. COVID vaccine boosters: the most important questions. Nature 2021;596:178–180. doi: 10.1038/d41586-021-02158-6.

8. Costa Clemens SA, Weckx L, Clemens R, Almeida Mendes AV, Ramos Souza A, Silveira MBV, et al. Heterologous versus homologous COVID-19 booster vaccination in previous recipients of two doses of CoronaVac COVID-19 vaccine in Brazil (RHH-001): a phase 4, non-inferiority, single blind, randomised study. Lancet 2022;399:521–529. doi: 10.1016/S0140-6736(22)00094-0.

9. Pérez-Then E, Lucas C, Monteiro VS, Miric M, Brache V, Cochon L, et al. Neutralizing antibodies against the SARS-CoV-2 Delta and Omicron variants following heterologous CoronaVac plus BNT162b2 booster vaccination. Nat Med 2022;28:481–485. doi: 10.1038/s41591-022-01705-6.

10. Cheng SMS, Mok CKP, Leung YWY, Ng SS, Chan KCK, Ko FW, et al. Neutralizing antibodies against the SARS-CoV-2 Omicron variant BA.1 following homologous and heterologous CoronaVac or BNT162b2 vaccination. Nat Med 2022;28:486–489. doi: 10.1038/s41591-022-01704-7.

11. Dolgin E. Omicron thwarts some of the world’s most-used COVID vaccines. Nature 2022;601:311. doi: 10.1038/d41586-022-00079-6.

12. McMenamin ME, Nealon J, Lin Y, Wong JY, Cheung JK, Lau EHY, et al. Vaccine effectiveness of one, two, and three doses of BNT162b2 and CoronaVac against COVID-19 in Hong Kong: a population-based observational study. Lancet Infect Dis 2022;22:1435–1443. doi: 10.1016/s1473-3099(22)00345-0.

13. Jara A, Undurraga EA, Zubizarreta JR, González C, Pizarro A, Acevedo J, et al. Effectiveness of homologous and heterologous booster doses for an inactivated SARS-CoV-2 vaccine: a large-scale prospective cohort study. Lancet Glob Health 2022;10:e798–e806. doi: 10.1016/s2214-109x(22)00112-7.

14. Yu X, Wei D, Xu W, Li Y, Li X, Zhang X, et al. Reduced sensitivity of SARS-CoV-2 Omicron variant to antibody neutralization elicited by booster vaccination. Cell Discov 2022;8:4. doi: 10.1038/s41421-022-00375-5.

15. Melo-González F, Méndez C, Peñaloza HF, Schultz BM, Piña-Iturbe A, Ríos M, et al. Humoral and cellular response induced by a second booster of an inactivated SARS-CoV-2 vaccine in adults. medRxiv 20222022.2008.2022.22279080. doi: 10.1101/2022.08.22.22279080.

16. Ranzani OT, Hitchings MDT, de Melo RL, de França GVA, Fernandes CdFR, Lind ML, et al. Effectiveness of an inactivated Covid-19 vaccine with homologous and heterologous boosters against Omicron in Brazil. Nature Communications 2022;13:5536. doi: 10.1038/s41467-022-33169-0.

17. Huang Z, Xu S, Liu J, Wu L, Qiu J, Wang N, et al. Effectiveness of inactivated and Ad5-nCoV COVID-19 vaccines against SARS-CoV-2 Omicron BA. 2 variant infection, severe illness, and death. BMC Medicine 2022;20:400. doi: 10.1186/s12916-022-02606-8.

18. Dai L, Gao GF. Viral targets for vaccines against COVID-19. Nat Rev Immunol 2021;21:73–82. doi: 10.1038/s41577-020-00480-0.

19. Li Y, Zhang Y, Zhou Y, Li Y, Xu J, Ai Y, et al. An RBD virus-like particle vaccine for SARS-CoV-2 induces cross-variant antibody responses in mice and macaques. Signal Transduct Target Ther 2023;8:173. doi: 10.1038/s41392-023-01425-4.

20. Bruun TUJ, Andersson AC, Draper SJ, Howarth M. Engineering a Rugged Nanoscaffold To Enhance Plug-and-Display Vaccination. ACS Nano 2018;12:8855–8866. doi: 10.1021/acsnano.8b02805.

21. Bachmann MF, Jennings GT. Vaccine delivery: a matter of size, geometry, kinetics and molecular patterns. Nat Rev Immunol 2010;10:787–796. doi: 10.1038/nri2868.

22. Lin YJ, Shih YJ, Chen CH, Fang CT. Aluminum salts as an adjuvant for pre-pandemic influenza vaccines: a meta-analysis. Sci Rep 2018;8:11460. doi: 10.1038/s41598-018-29858-w.

23. Sahin U, Muik A, Derhovanessian E, Vogler I, Kranz LM, Vormehr M, et al. COVID-19 vaccine BNT162b1 elicits human antibody and T(H)1 T cell responses. Nature 2020;586:594–599. doi: 10.1038/s41586-020-2814-7.

24. Song JY, Choi WS, Heo JY, Lee JS, Jung DS, Kim SW, et al. Safety and immunogenicity of a SARS-CoV-2 recombinant protein nanoparticle vaccine (GBP510) adjuvanted with AS03: A randomised, placebo-controlled, observer-blinded phase 1/2 trial. EClinicalMedicine 2022;51:101569. doi: 10.1016/j.eclinm.2022.101569.

25. Ward BJ, Gobeil P, Seguin A, Atkins J, Boulay I, Charbonneau PY, et al. Phase 1 randomized trial of a plant-derived virus-like particle vaccine for COVID-19. Nat Med 2021;27:1071–1078. doi: 10.1038/s41591-021-01370-1.

26. Zhang Z, He Q, Zhao W, Li Y, Yang J, Hu Z, et al. A Heterologous V-01 or Variant-Matched Bivalent V-01D-351 Booster following Primary Series of Inactivated Vaccine Enhances the Neutralizing Capacity against SARS-CoV-2 Delta and Omicron Strains. J Clin Med 2022;11. doi: 10.3390/jcm11144164.

27. Ai J, Wang X, He X, Zhao X, Zhang Y, Jiang Y, et al. Antibody evasion of SARS-CoV-2 Omicron BA.1, BA.1.1, BA.2, and BA.3 sub-lineages. Cell Host Microbe 2022;30:1077–1083 e1074. doi: 10.1016/j.chom.2022.05.001.

28. Nilles EJ, Paulino CT, de St Aubin M, Duke W, Jarolim P, Sanchez IM, et al. Tracking immune correlates of protection for emerging SARS-CoV-2 variants. Lancet Infect Dis 2023;23:153–154. doi: 10.1016/s1473-3099(23)00001-4.

29. Lang-Meli J, Luxenburger H, Wild K, Karl V, Oberhardt V, Salimi Alizei E, et al. SARS-CoV-2-specific T-cell epitope repertoire in convalescent and mRNA-vaccinated individuals. Nature Microbiology 2022;7:675–679. doi: 10.1038/s41564-022-01106-y.

30. Keeton R, Tincho MB, Ngomti A, Baguma R, Benede N, Suzuki A, et al. Author Correction: T cell responses to SARS-CoV-2 spike cross-recognize Omicron. Nature 2022;604:E25. doi: 10.1038/s41586-022-04708-y.

31. Assawakosri S, Kanokudom S, Suntronwong N, Auphimai C, Nilyanimit P, Vichaiwattana P, et al. Neutralizing Activities against the Omicron Variant after a Heterologous Booster in Healthy Adults Receiving Two Doses of CoronaVac Vaccination. medRxiv 20222022.2001.2028.22269986. doi: 10.1101/2022.01.28.22269986.

32. Filardi BA, Monteiro VS, Schwartzmann PV, do Prado Martins V, Zucca LER, Baiocchi GC, et al. Age-dependent impairment in antibody responses elicited by a homologous CoronaVac booster dose. Sci Transl Med 2023;15:eade6023. doi: 10.1126/scitranslmed.ade6023.

33. Zhu FC, Guan XH, Li YH, Huang JY, Jiang T, Hou LH, et al. Immunogenicity and safety of a recombinant adenovirus type-5-vectored COVID-19 vaccine in healthy adults aged 18 years or older: a randomised, double-blind, placebo-controlled, phase 2 trial. Lancet 2020;396:479–488. doi: 10.1016/s0140-6736(20)31605-6.

34. Grifoni A, Weiskopf D, Ramirez SI, Mateus J, Dan JM, Moderbacher CR, et al. Targets of T Cell Responses to SARS-CoV-2 Coronavirus in Humans with COVID-19 Disease and Unexposed Individuals. Cell 2020;181:1489–1501 e1415. doi: 10.1016/j.cell.2020.05.015.

35. Sette A, Crotty S. Pre-existing immunity to SARS-CoV-2: the knowns and unknowns. Nat Rev Immunol 2020;20:457–458. doi: 10.1038/s41577-020-0389-z.

