## Supplementary material for "Safety and immunogenicity of a heterologous booster with an RBD virus-like particle vaccine following two- or three-dose inactivated COVID-19 vaccine": Table S1, Table S2, Figure S1 and Supplementary Methods

#### Table S1 Pseudovirus neutralizing antibody GMT, SCR, GMFR against SARS-CoV-2 at baseline and 14, 28, 90 days after booster

| Days after booster | Prototype | | | | Omicron BA.4/5 | | | |
| --- | --- | --- | --- | --- | --- | --- | --- | --- |
|  | Booster after two-dose primary series of ICV | | | Booster after three-dose ICV | Booster after two-dose primary series of ICV | | | Booster after three-dose ICV |
|  | 30µgLYB001 | 60µgLYB001 | CoronaVac | 30µgLYB001 | 30µgLYB001 | 60µgLYB001 | CoronaVac | 30µgLYB001 |
| Day 0 | | | | | | | | |
| n | 30 | 30 | 29 | 30 | 30 | 30 | 29 | 30 |
| GMT | 8.5 (6.3, 11.6) | 7.0 (5.2, 9.2) | 9.0 (6.3, 12.7) | 83.9 (55.5, 126.8) | 5.0 (5.0, 5.0) | 5.0 (5.0, 5.0) | 5.0 (5.0, 5.0) | 9.4 (6.2, 14.0) |
| SCR | NA | NA | NA | NA | NA | NA | NA | NA |
| GMFR | NA | NA | NA | NA | NA | NA | NA | NA |
| Day 14 | | | | | | | | |
| n | 30 | 30 | 29 | 29 | 30 | 30 | 29 | 29 |
| GMT | 771.6 (452.3, 1316.5) | 522.8 (334.5, 817.0) | 198.2 (122.5, 320.8) | 1124.4 (727.9, 1737.1) | 109.6 (52.3, 229.6) | 67.3 (36.5, 124.0) | 29.9 (17.4, 51.4) | 135.7 (75.6, 243.5) |
| SCR | 100.0 (88.4, 100.0) | 100.0 (88.4, 100.0) | 86.2 (68.3, 96.1) | 89.7 (72.6, 97.8) | 86.7 (69.3, 96.2) | 86.7 (69.3, 96.2) | 75.9 (56.5, 89.7) | 93.1 (77.2, 99.2) |
| GMFR | 90.4 (51.9, 157.4) | 75.3 (48.3, 117.4) | 22.1 (12.8, 38.1) | 13.6 (9.4, 19.7) | 21.9 (10.4, 45.9) | 13.4 (7.3, 24.8) | 6.0 (3.5, 10.3) | 15.0 (9.1, 24.6) |
| Day 28 | | | | | | | | |
| n | 30 | 30 | 29 | 30 | 30 | 30 | 29 | 30 |
| GMT | 1237.8 (747.2, 2050.6) | 554.3 (374.6, 820.2) | 181.9 (107.6, 307.6) | 1200.2 (831.5, 1732.3) | 201.1 (102.7, 393.7) | 63.0 (35.1, 113.1) | 29.2 (16.9, 50.3) | 115.3 (63.9, 208.1) |
| SCR | 100.0 (88.4, 100.0) | 100.0 (88.4, 100.0) | 82.8 (64.2, 94.2) | 90.0 (73.5, 97.9) | 93.3 (77.9, 99.2) | 86.7 (69.3, 96.2) | 75.9 (56.5, 89.7) | 83.3 (65.3, 94.4) |
| GMFR | 145.0 (85.6, 245.7) | 79.8 (53.3, 119.4) | 20.3 (11.6, 35.6) | 14.3 (10.2, 20.1) | 40.2 (20.6, 78.7) | 12.6 (7.0, 22.6) | 5.8 (3.4, 10.1) | 12.3 (7.6, 20.0) |
| Day 90 | | | | | | | | |
| n | 30 | 30 | 29 | 30 | 30 | 30 | 29 | 30 |
| GMT | 384.3 (232.4, 635.5) | 336.8 (215.5, 526.3) | 107.7 (62.2, 186.2) | 609.1 (437.3, 848.4) | 44.4 (23.2, 85.2) | 32.2 (18.1, 57.7) | 14.1 (8.8, 22.5) | 42.0 (24.7， 71.4) |
| SCR | 100.0 (88.4, 100.0) | 100.0 (88.4, 100.0) | 85.2 (66.3, 95.8) | 80.0 (61.4, 92.3) | 80.0 (61.4, 92.3) | 70.0 (50.6, 85.3) | 55.6 (35.3, 74.5) | 66.7 (47.2, 82.7) |
| GMFR | 45.0 (27.1, 74.8) | 48.5 (30.2, 77.9) | 11.9 (6.5, 21.8) | 7.3 (5.3, 9.9) | 8.9 (4.6, 17.0) | 6.5 (3.6, 11.5) | 2.8 (1.8, 4.5) | 4.5 (3.1, 6.5) |

*GMT, SCR, GMFR are presented as n (95%CI). GMT: geometric mean titer, SCR: seroconversion rate, GMFR: geometric mean fold rise.*

#### Table S2 RBD-specific IFN-γ, IL-2, IL-4 secreting T cells measured by ELISpot assay

|  | SFCs/ 3×10^5^PBMCs | | | | |
| --- | --- | --- | --- | --- | --- |
|  | Booster after two-dose primary series of ICV | | | | Booster after three-dose ICV |
|  | 30µg LYB001 | 60µg LYB001 | CoronaVac | | 30µg LYB001 |
| Day 0 | | | | | |
| IFN-γ | 1.0 (0.0, 2.0) | 0.0 (0.0, 1.0) | | 0.0 (0.0, 2.0) | 0.0 (0.0, 1.0) |
| IL-2 | 4.0 (1.0, 10.0) | 2.5 (0.0, 5.0) | | 5.0 (2.0, 8.0) | 0.0 (0.0, 3.0) |
| IL-4 | 1.0 (0.0, 3.0) | 0.0 (0.0, 1.0) | | 1.0 (0.0, 1.0) | 0.0 (0.0, 2.0) |
| Day 14 | | | | |  |
| IFN-γ | 23.0 (8.0, 68.0) | 23.0 (10.0, 42.0) | | 2.0 (0.0, 4.0) | 18.0 (5.0, 49) |
| IL-2 | 48.0 (26.0, 145.0) | 39.0 (21.0, 70.0) | | 5.0 (3.0, 8.0) | 54.0 (30.0, 99.0) |
| IL-4 | 12.0 (4.0, 36.0) | 8.0 (4.0, 33.0) | | 1.0 (0.0, 1.0) | 18.0 (7.0, 43.0) |

*Data is presented median (Q1, Q3). RBD: receptor binding domain, SFCs: spot forming cells*

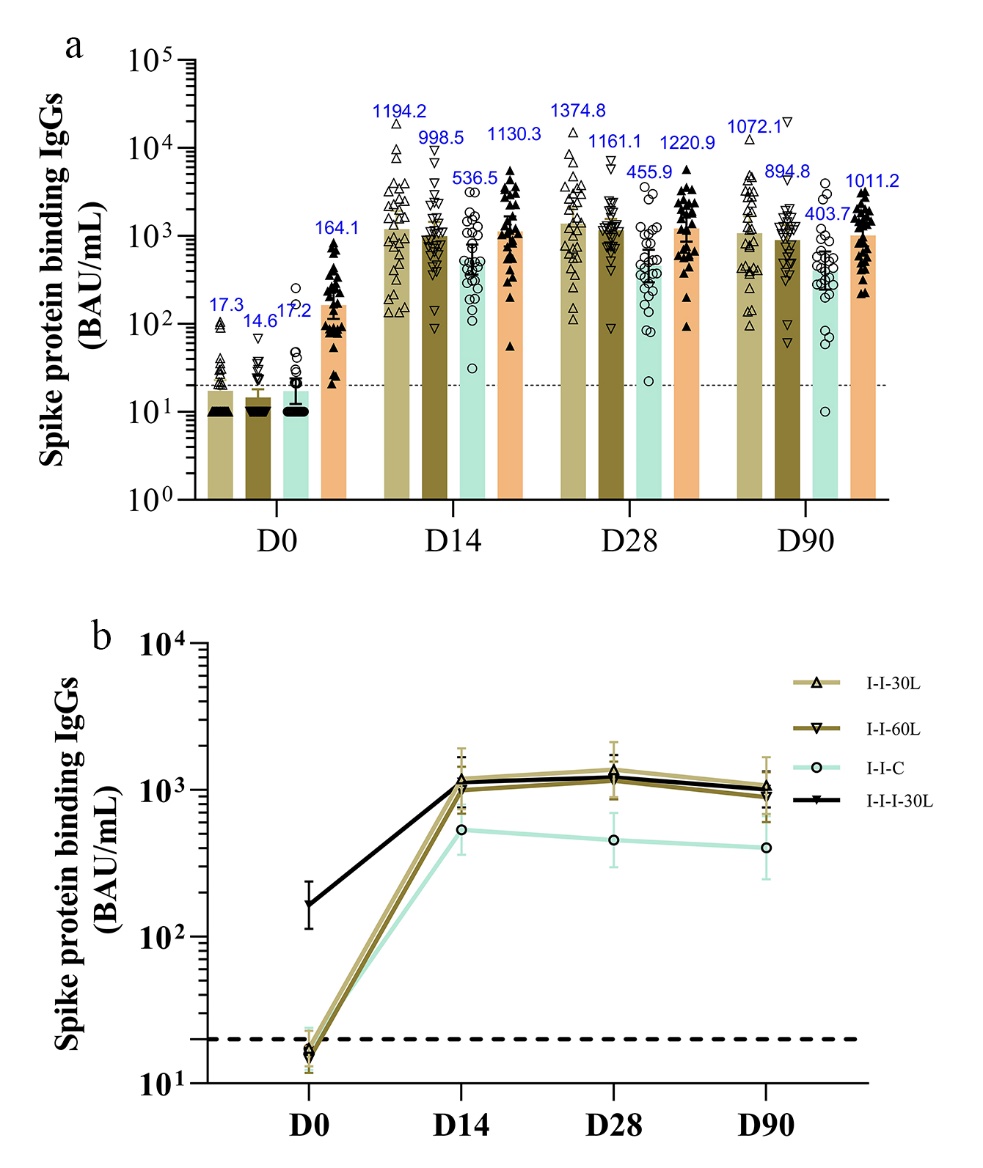

***Figure S1 The spike protein binding IgGs at baseline and 14, 28, 90 days after booster.***

*Antibody values reported as below the lower limit of detection (LOD=20) were replaced by 0.5*LOD. The data in the I-I-30L, I-I-60L, I-I-C, and I-I-I-30L groups are indicated by* △, ▽, ○, ▲*, respectively. I-I-30L: 30μg LYB001 booster after two-dose inactivated COVID-19 vaccine, I-I-60L: 60μg LYB001 booster after two-dose inactivated COVID-19 vaccine, I-I-C: CoronaVac booster after two-dose inactivated COVID-19 vaccine, I-I-I-30L: 30μg LYB001 booster after three-dose inactivated COVID-19 vaccine.*

### Supplementary methods

#### Pseudovirus neutralization assay

Pseudovirus neutralization assay was performed using the VSV-based SARS-CoV-2 pseudovirus bearing Spike protein of Prototype, Omicron BA.4/5, which were provided by Zhongxingrongchuang (Beijing) Bio-Tech Co. Ltd. The SARS-CoV-2 pseudotyped virus neutralization test began with 3-fold serially diluted heat-inactivated serum samples, which started at appropriate dilution levels determined by the GMC of Spike protein antibody and then mixed with a certain amount diluted SARS-CoV-2 pseudovirus for about 1h at 37 °C with 5% CO_2_ supplementation. Then the mixture was added with Vero cells (2×10^4^ per well). After that, the target cells were incubated for about 24 h at 37 °C with 5% CO_2_ supplementation, and the amount of pseudovirus entering the target cells was calculated by detecting the expression of luciferin using Bio-Lite Luciferase Assay System, supplied by Vazyme Biotech Co., Ltd. The 50% neutralization titer (pVNT_50_) was calculated and was defined as the reciprocal of serum dilution at which the relative light units (RLU) were reduced by 50% compared with the virus control wells. Samples with values ≥ 10 were defined as seropositive.

#### ELISpot assay

T cell responses were assessed by ELISpot assay using Human IFN-γ, IL-2 and IL-4 ELISPOT^Pro^ kit (Mabtech) following the manufacturer’s instructions. Plates were washed with sterile PBS (Biotopped) and blocked with culture medium containing 10% FBS (ExCellBio). The thawed PBMCs were recovered, and re-suspended to seed on the plates with a final concentration of 3×10^5^/well. Negative controls comprised culture medium with 1%DMSO and 1% penicillin-streptomycin, and positive controls comprised anti-CD3 mAb CD3-2 with 2 μg/mL phorbol myristate acetate (PMA) and 4 μg/mL ionomycin (ION). As standard, 3×10^5^ cells per well were stimulated in triplicate with 2μg/ml overlapping peptide pools covering the full sequence of receptor binding domain (RBD, aa 319-532) from prototype SARS-CoV-2 for 21h±30min at 37 °C with 5% CO_2_ supplementation, and a total of 21 peptides were used. After washes (5 times with PBS), 100μl diluted 7-B6-HRP (horseradish peroxidase-conjugated detection mAb 7-B6-1 1:200 in filtered PBS containing 0.5% FBS) was added to the plates and incubated for 2 h±5min at room temperature, then after another round of washes, 100μl BCIP/NBT-plus substrate was added and incubated for 10±1min  min at room temperature. Then the plate was rinsed, left to air dry, then underwent spot counting using an AID ELISpot Reader System. Mean spot counts for negative control wells were subtracted from the mean of test wells to generate normalized readings, these are presented as spot forming cells (SFCs) per 3×10^5^ PBMCs. The value of (median+2×SD) was used as the lower limit to indicate a positive response in the test cohort.
